# Quality, consistency, and clinical safety of AI-generated versus clinician-written clinical notes: a multi-country paired simulation study

**DOI:** 10.64898/2026.08.18.26360701

**Authors:** Henry Bergman, Vivian Liu, Ben Austin, Sarah Ali, Elliot Taylor, Merle Fiedler, Celene Sandiford, Rebecca Blanchard, Carlos Casanovas, Victor Najar, Giulia Pedrazzini, Theo Markopouliotis, Ferdinand Vermersch

**Author notes:** **Corresponding author:** Henry Bergman.

## Abstract

**Background:** Ambient AI documentation tools (“scribes”) are entering routine clinical practice at scale, but the evidence comparing the notes they produce against clinician-written notes is dominated by single-site, single-language studies that rely on human review to find errors — a method known to miss most documentation errors.

**Methods:** We conducted a paired simulation across five countries and languages (Cambridge/English, Barcelona/Spanish, Milan/Italian, Paris/French, Cologne/German; 385 paired consultations, 770 notes). From each actor-performed consultation, an AI scribe (Heidi) and a junior-to-middle-grade clinician independently produced a note. Notes were scored on the PDQI-9 by evaluators blinded to authorship. Documentation errors were identified by two methods of deliberately different sensitivity — clinician adjudication, and a calibrated automated reviewer externally validated against a blinded ten-clinician panel — then graded for clinical risk by a three-model panel. The co-primary outcomes were PDQI-9 total and Critical+High error burden, the latter reported under both detection arms. The analysis plan was registered before any pooling across sites.

**Results:** AI notes scored higher than clinician notes on the PDQI-9 (40.6 vs 35.6; difference +5.08, 95% CI 4.6–5.6; Cohen’s dz=0.55), consistently across all five sites (dz 0.41–0.75), and were less dispersed (5.7% of AI vs 27.8% of clinician notes fell below the study pre-specified low-score threshold (<32)). On the principal safety outcome — the paired probability that a note carried ≥1 Critical+High error — clinician notes were affected more often under both detection arms: 61.0% versus 24.4% by the calibrated reviewer (relative risk 2.50, 95% CI 2.09–3.00) and 21.8% versus 6.2% by clinician adjudication (relative risk 3.50, 95% CI 2.32–5.27). The difference was largest for omissions. Unaided clinician review identified roughly 12% of the errors the calibrated reviewer retained, and a smaller fraction in AI notes than in clinician notes.

**Conclusions:** In this simulation, AI-generated notes scored higher on documentation quality, varied less, and carried fewer clinically significant errors than notes written on the same consultations by junior-to-middle-grade clinicians. The magnitude of the safety difference depends on the sensitivity of error detection, so we report both detection regimes and bound rather than point-estimate the absolute error rate. Extension to live practice, consultant-authored documentation, and notes as filed after clinician editing remains to be established.

**One sentence description:** In a five-country, five-language paired simulation, AI-generated clinical notes scored higher on documentation quality and carried fewer clinically significant errors than notes written on the same consultations by junior-to-middle-grade clinicians.

## Introduction

Clinical documentation is a substantial, error-prone component of clinical work.^1^ Ambient AI scribes, which generate a structured note from the consultation audio, are now being adopted at scale with the explicit promise of reducing documentation burden while maintaining or improving note quality.^2-4^ The appropriate benchmark for such a tool is not an abstract standard of perfection but the clinician note it replaces, and that comparison has to be made on the same consultation: encounters vary enormously in complexity, and the AI and clinician notes of a single encounter are naturally paired. The comparator also has to be a specific clinical population. The bulk of routine clinical documentation (triage, admission, clinic and ward-round notes) is produced not by consultants but by junior-to-middle-grade clinicians, by far the highest-volume note-writers in most health systems.^5-7^ We therefore deliberately sampled this primary documentation workforce as the most clinically relevant point of comparison for a scribe intended to support exactly that work.

Two methodological problems limit the existing evidence. First, most studies are single-site and single-language, leaving open whether any advantage generalises across health systems and, critically, across languages, a first-order question for a globally deployed tool.^8,9^ Second, and more subtly, documentation errors are usually identified by human reviewers, who are known to miss a large fraction of the errors actually present.^10^ If human review is insensitive, reported error rates reflect the detector as much as the notes, and any comparison between AI and clinician notes inherits that insensitivity. A study that wishes to make a credible safety claim must therefore either use a more sensitive, calibrated detector, or at minimum measure how detection sensitivity changes the answer.

We address both problems directly. We ran a paired simulation across five countries and languages in which an AI scribe and a clinician each documented the same simulated consultation, and we identified errors with two independent methods of deliberately different sensitivity, clinician adjudication and a calibrated automated reviewer, so that the detection-method dependence of the result is itself a measured outcome rather than a hidden assumption. Every error was graded for clinical risk by an independent multi-model panel. The full analysis, including all supplementary and robustness analyses, was pre-specified in a statistical analysis plan frozen before any pooling across sites, and is reported in the Supplementary Appendix.

## Methods

### Design, Cohort, and Consultations

This was a prospective, paired, multi-country simulation study. From each simulated consultation, two clinical notes were produced independently and from the same source material: one generated by the AI scribe (Heidi) and one written by a clinician. Because both notes derive from the identical consultation, every AI–clinician comparison is naturally paired by session, removing between-encounter variation in case complexity from the primary contrast.

Five sites were analysed, each corresponding to a region and language: Cambridge, United Kingdom (English); Paris, France (French); Cologne, Germany (German); Milan, Italy (Italian); and Barcelona, Spain (Spanish). Of 400 consultations across these five sites, 385 paired consultations (770 notes) contributed to the pooled analysis (Cambridge n=80, Paris 80, Milan 80, Barcelona 79, Cologne 66). Fifteen consultations were excluded because one of the two primary note evaluations was missing following adjudicator non-completion, principally in Cologne (14 sessions) and Barcelona (1 session); exclusions followed the pre-specified requirement for a complete AI-clinician pair. A separate 15-consultation Allied Health arm was excluded from the main analysis before pooling because it lacked the inter-rater reliability cross-over substudy required for the primary quality outcome, and used a structurally different note type. Its descriptive results are reported in Supplementary Table S8.

**Figure 1.**
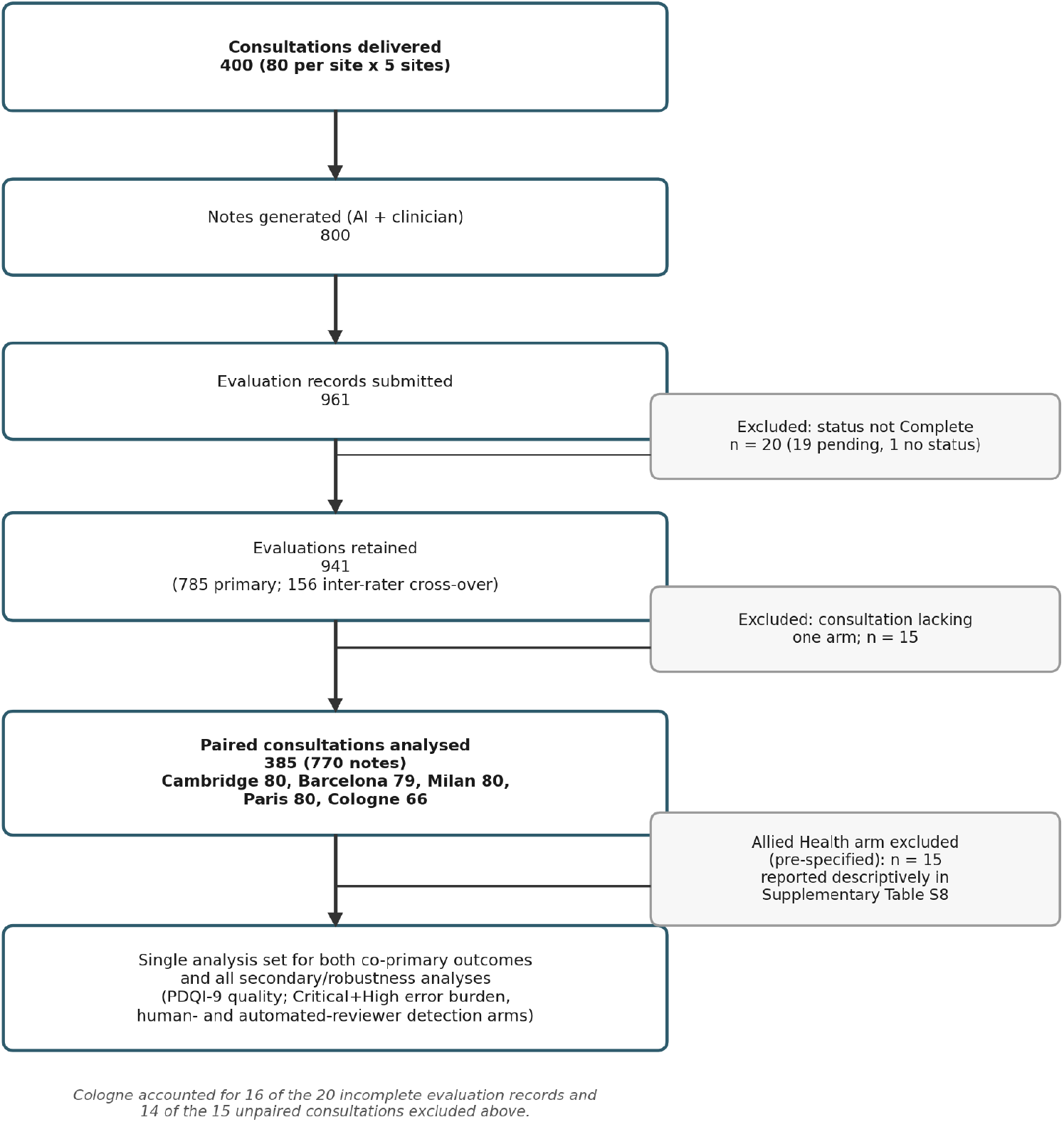
Study flow diagram. Counts reflect the frozen analysis dataset.

Consultations were performed by trained actors from controlled clinical scripts derived from real encounters, so that the ground-truth content of each encounter was known in advance, a prerequisite for unambiguous error adjudication that is not achievable with live recordings. Scripts were deliberately seeded with naturalistic complexity, speech dysfluencies, patient-led tangents, hedging and diagnostic uncertainty, and were recorded under four acoustic conditions: clean audio and background noise at 40, 50 and 60 dB. Each note-writing clinician contributed multiple sessions (approximately 10), enabling estimation of within-clinician consistency.

We assessed the AI’s unedited first-draft note against the clinician’s note. This was a deliberate, pre-specified choice: the unedited draft is the artefact over which the system itself has full control, so assessing it sets a floor on performance that is independent of downstream editing. The two documenters also did not have equivalent access to the source material. The AI operated on a persistent transcript; the clinician documented from a single live hearing, with no replay and a maximum of 10 minutes to complete the note,where prospectively measured time in notes averages approximately 10.3 minutes per outpatient appointment before any ambient documentation support11, and it therefore favours the clinician arm. We did not equalise this asymmetry, because persistent capture is the mechanism by which an ambient scribe adds value, and removing it would evaluate a system that does not exist. Full detail on scripts, participants, and documentation conditions is in the Supplementary Appendix.

### Participants

Note-writing clinicians (8 per site, 40 in total) were junior-to-middle-grade doctors spanning Foundation Year to Specialty Registrar or equivalent (mean age 29 years, range 24–34). This grade distribution was intentional, for the reason given above.^5-7^ Blinded clinical evaluators (67 unique) scored notes without knowing their authorship. Note-writers were recruited openly from doctors currently working and registered to practise in the jurisdiction of their site, and documenting in that site’s language, which reproduces the documentation workforce as it actually exists rather than a filtered subset of native speakers. Full cohort and participant detail, by site, is reported in Supplementary Table S1.

### Ethics and Consent

All participants, actors, note-writing clinicians, and evaluators provided written informed consent to participate and to the research use of the resulting notes and recordings. No real patients participated and no patient-identifiable data were used: consultation scripts were thematically derived from real production clinician encounter topics to model clinical realism, but reused none of the original identifying detail. Scripts were therefore entirely synthetic, non-reidentifiable to the inspiring sessions, and created within an appropriate governance framework. For the United Kingdom site, the Health Research Authority decision tool returned a determination that NHS Research Ethics Committee review was not required.^12^

### Quality Outcome and Error Identification

Note quality was measured with the PDQI-9 (Physician Documentation Quality Instrument: nine items, each scored 1–5, giving a total of 9–45; the items are up-to-date, accurate, thorough, useful, organised, comprehensible, succinct, synthesized, and internally consistent).^13^ The instrument and evaluator instructions were professionally translated into French, German, Italian, and Spanish for the non-English sites, with independent back-translation and reconciliation, following established guidance for the cross-cultural adaptation of self-report instruments^14^. Each note was scored by blinded clinical evaluators, with a pre-specified 20% inter-rater cross-over. Because no validated low-quality cut-point for PDQI-9 has been published in any of these five languages to our knowledge, the registered analysis plan defined a low-scoring note as one falling below the 25th percentile of the pooled PDQI-9 distribution across both note types; in the frozen analysis dataset this corresponded to a score <32.The two co-primary outcomes were PDQI-9 total score and Critical+High error burden; the latter is reported separately under each of the two error-detection arms described below.

Documentation errors were defined in two classes, following standard practice for generated clinical text.^15,16^ Commission errors are statements in the note that are not supported by, or that contradict, the consultation. Omission errors are clinically relevant content present in the consultation but absent from the note. Errors were identified by two independent methods of differing sensitivity. In the human-identification arm, clinician adjudicators manually flagged perceived omission and commission errors in free text. In the calibrated automated-reviewer arm, an LLM judge^17^ reviewed every note blind to authorship, and an independent LLM screen classified each candidate as a genuine error or a defensible non-error (faithful paraphrase, reasonable inference, or appropriate clinical concision), applying the same criterion identically to both arms. The judge and screen belong to a different model family from the note-generation system under evaluation, so self-preference bias, in which an LLM evaluator favors text produced by its own family,^18^ cannot operate through shared lineage at those stages.

### External Validation and Severity Grading

The automated reviewer’s false-positive screen and severity panel were validated in a separate, pre-registered, blinded human-adjudication substudy.^19^ Ten external clinicians, independent of the sponsor and study team, adjudicated a stratified sample of 434 pipeline flags blinded, to note authorship, identification source, and the pipeline’s own verdict. They were also independent of one another, adjudicating separately and without sight of any other adjudicator’s judgements, and they were remunerated at a fixed sessional rate that did not depend on their findings. Because no human gold standard for documentation error exists (inter-clinician agreement on genuine-error status was only fair, Gwet’s AC1 0.24), validation was framed as fitness for purpose rather than accuracy: judge–clinician agreement (64%) fell within the range of clinician–clinician agreement (59%), and the instrument behaved near-symmetrically across AI and clinician notes. The one asymmetry we detected runs against the direction of our reported result, and is quantified in the Discussion. A Bayesian latent-class triangulation (Hui–Walter paradigm)^20^ placed pipeline sensitivity at 0.83 and specificity at 0.87. Full validation results are reported in Supplementary Tables S3 and S6.

Every identified error was graded for clinical risk on a severity × likelihood matrix (each 1–5), using a rubric aligned with ISO 14971 medical-device risk management^21^ and UK health-IT clinical risk management standards.^22^ Grading used an independent three-model panel (GPT-5.5, Claude Opus 4.8, Gemini 2.5 Pro), with consensus taken by majority vote. Tiers were Critical (16–25), High (10–15), Moderate (5–9) and Low (1–4). The pre-specified primary safety summaries were Critical+High density (severe errors per note) and burden (percentage of notes with at least one severe error).

### Statistical Analysis

This study is reported in accordance with the TRIPOD-LLM reporting guideline for studies using large language models.^23^ The primary quality comparison used a paired Wilcoxon signed-rank test on session-level PDQI-9, with Cohen’s dz^21^ and 95% CI, run per site and pooled. The study was designed around a fixed operational protocol rather than a prospective sample-size calculation. The pre-registered, pre-unblinding power assessment assumed an unpaired effect size of 0.3 to 0.5 and a within-pair correlation of 0.3 to 0.5; under the most conservative scenario (d = 0.3, correlation 0.3) the planned pooled n = 400 provided greater than 99% approximate power while per-site n = 80 provided 61%, and the minimum detectable paired effect at 80% power was dz = 0.31 per site and dz = 0.14 pooled. The realised pooled sample of 385 was therefore materially unchanged for the primary pooled comparison. Single-site subgroup analyses (n of approximately 20) were powered at 20% to 27% for small effects and were pre-specified as exploratory, without inferential correction. Consistency was assessed with Levene’s test,^25^ the coefficient of variation, and the proportion of notes falling below the pre-specified low-score threshold. Item-level analysis applied paired Wilcoxon to each of the nine PDQI items with Benjamini–Hochberg FDR correction.^26^ Safety differentials used paired McNemar tests on burden together with paired comparison of density; site-adjusted negative-binomial GLMs were pre-specified as a supporting analysis and are not reported on the final analysis set. A Bayesian crossed-random-effects model (bambi^27^/PyMC^28^), PDQI ~(1|session) + (1|evaluator) + (1|notewriter), corroborated the primary contrast (Supplementary Table S9). Replication across sites was summarised with the I^2^ heterogeneity statistic^29^ and a forest plot. Clinical significance used a distribution-based MCID of half the standard deviation of the pooled score distribution.^30^ Analyses were implemented in Python (pandas,^31^ scipy,^32^ statsmodels,^33^ pingouin^34^). The statistical analysis plan was registered on the Open Science Framework before any pooling across sites(https://osf.io/j46vz/overview; doi:10.17605/OSF.IO/J46VZ); the registered plan is reproduced as Supplementary File S1, and departures from it are itemised in Supplementary Table S2.

Sample size and power were fixed in the registered analysis plan before any data were unblinded. The study was powered on the primxary quality contrast, with the session as the unit of analysis and 80 paired consultations planned per site (400 pooled). Power was estimated for the paired t-test as a conservative approximation to the paired Wilcoxon signed-rank test, whose asymptotic relative efficiency is at least 0.955 for approximately normal distributions and can exceed it under skew. The planning assumption was an unpaired Cohen’s d of 0.5, corresponding to a PDQI-9 difference of approximately 3 points at a pooled SD of 6, with a within-session correlation between paired notes of 0.3, giving a paired dz of 0.42. Under those assumptions the per-site analysis had 96% power and the pooled analysis exceeded 99%, at a two-sided alpha of 0.05. Under a deliberately conservative scenario (d = 0.3, correlation 0.3, dz = 0.25) per-site power fell to 61% while pooled power remained above 99%. The minimum detectable paired effect at 80% power was dz = 0.31 per site and dz = 0.14 pooled. Subgroup analyses pooled across sites (n of approximately 100) were powered at 85% or above under conservative assumptions; single-site subgroup analyses (n of approximately 20) were powered at only 20% to 27% for small effects and were therefore pre-specified as exploratory, are not subject to inferential correction, and are reported as directional only. The observed pooled effect (dz = 0.55) exceeds the primary planning assumption, and per-site null results, had any occurred, would have been interpreted as inconclusive and not as evidence of absence.

### Bias Mitigation and Validation

All authors are employees of Heidi Health, the developer of the AI scribe and the automated reviewer evaluated here; this is a sponsor-conducted evaluation. Several design features constrain, but do not eliminate, the scope for directional bias. The error-identification judge and false-positive screen, the stages that determine error counts, are from a different model family than the note-generation system, so self-preference bias cannot operate through shared lineage there, and the severity-grading panel spans three model families. The pipeline is blind to authorship throughout, and the clinician-worse safety result is present in the raw, pre-calibration judge output. The external adjudication panel that validated the instrument was independent of the sponsor and of its members, as specified above, and was remunerated irrespective of its findings. Most importantly, the largest effect, the omission difference, is independently reproduced by the human-adjudication arm, which uses none of these tools. The full set of blinding, non-discrimination, and external-validation checks is reported in Supplementary Tables S3 and S6.

## Results

### Note Quality

AI notes scored higher than clinician notes on the PDQI-9 total: pooled mean 40.6 versus 35.6, a difference of +5.08 points (95% CI 4.6–5.6; Cohen’s dz=0.55, 95% CI 0.46–0.66), in the same direction at all five sites (Table 1). On the 9–45 scale this is approximately 14% of the instrument’s 36-point range. At the individual-consultation level, the AI note scored higher in 66.8% of the 385 pairs, equal in 7.5%, and lower in 25.7% (Supplementary Figure S1). AI notes were approximately twice as long as clinician notes (mean 1,645 versus 797 characters), but in a linear model adjusting for note length the estimated difference moved only from +5.05 to +4.95 points, and length was not independently associated with PDQI-9 score in that model, so the difference is not attributable to length alone.

**Table 1.** Per-site PDQI-9: AI advantage replicates across all five languages.

| Site | N | AI, mean | Clinician, mean | Difference | Cohen's $d_z$ |
| --- | --- | --- | --- | --- | --- |
| Cambridge (English) | 80 | 40.8 | 37.1 | +3.61 | 0.49 |
| Paris (French) | 80 | 39.2 | 35.1 | +4.19 | 0.41 |
| Milan (Italian) | 80 | 41.2 | 36.5 | +4.73 | 0.60 |
| Barcelona (Spanish) | 79 | 42.1 | 37.7 | +4.44 | 0.60 |
| Cologne (German) | 66 | 39.7 | 30.6 | +9.14 | 0.75 |
| Pooled | 385 | 40.6 | 35.6 | +5.08 | 0.55 |

PDQI-9: Physician Documentation Quality Instrument, range 9–45, higher = better documentation quality.<sup>11</sup> Wilcoxon $p<0.001$ at every site and pooled (pooled $p=1.4\times 10^{-21}$ ). Differences are computed from unrounded means and may therefore differ slightly from the difference of the rounded means shown. Source: authors' analysis of the 385-consultation paired dataset.

AI scores were also less dispersed than clinician scores (SD 4.97 versus 7.76, Levene p=4×10^−13^), with 5.7% of AI versus 27.8% of clinician notes falling below a pre-specified low-score threshold (the lowest quartile of the pooled PDQI-9 distribution, corresponding to a score below 32), a 4.9-fold difference in the proportion of low-scoring notes (Figure 2; Supplementary Figures S2 and S3). All nine PDQI-9 items differed in favour of the AI note after FDR correction; the largest differences were on Thorough, Accurate and Organized, and the smallest on Succinct, consistent with the length and safety findings (Supplementary Figure S4). The direction of the difference was consistent across clinical settings and noise conditions, and a Bayesian crossed-random-effects model accounting for session, evaluator and note-writer estimated the clinician–AI difference at −5.16 PDQI points (94% credible interval −5.90 to −4.42; Supplementary Table S9 and Figure S5). Consistency, however, was asymmetric between the arms: the AI mean varied little across the five health systems (39.2–42.1) whereas the clinician mean varied 2.4-fold more widely (30.6–37.7), driven mainly by a lower clinician baseline at Cologne (Supplementary Figure S10). Between-site heterogeneity in the site mean differences was correspondingly substantial (I^2^=62.6%); the AI advantage was nonetheless positive and significant at every site taken individually, and excluding Cologne the site differences ranged from +3.61 to +4.73 points.

**Figure 2.**
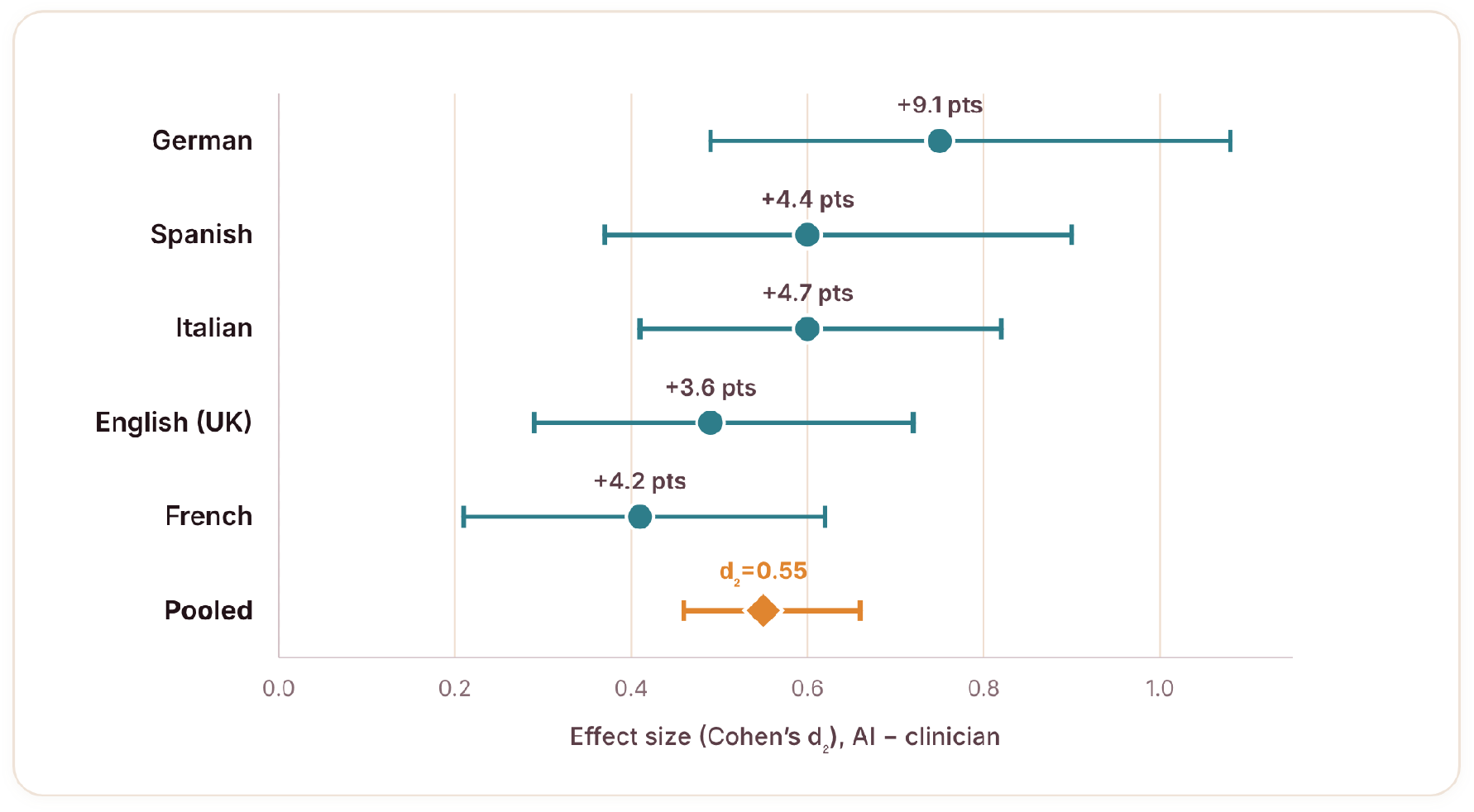
AI note-quality advantage (PDQI-9) by site. Paired effect size (Cohen’s dz), AI minus clinician, with 95% CI. PDQI-9 range 9–45, higher = better documentation quality. The advantage is positive and significant at every site; the diamond is the pooled estimate. Source: authors’ analysis of the 385-consultation paired dataset.

### Clinically Significant Errors

The pre-specified principal safety comparison is the paired probability that a note carries at least one Critical+High error, reported separately under each detection arm. Under the calibrated reviewer, 94 of 385 AI notes (24.4%) and 235 of 385 clinician notes (61.0%) carried such an error, a paired risk difference of +36.6 percentage points (95% CI +30.7 to +42.6; relative risk 2.50, 95% CI 2.09–3.00). Under clinician adjudication, the comparison gave 24 of 385 (6.2%) versus 84 of 385 (21.8%), relative risk 3.50 (95% CI 2.32–5.27). The direction and significance of the principal outcome do not depend on which detector is used, although the absolute rates differ substantially between arms (Table 2).

**Table 2.** Critical+High error density by detection arm.

| Error type | Detection arm | AI, per note | Clinician, per note | Clinician:AI ratio |
| --- | --- | --- | --- | --- |
| Commission | Calibrated automated reviewer | 0.24 | 0.58 | 2.4x |
| Omission | Calibrated automated reviewer | 0.094 | 0.683 | 7.3x |
| Commission | Clinician adjudication (FP-stripped) | 0.042 | 0.078 | 1.9x |
| Omission | Clinician adjudication (FP-stripped) | 0.039 | 0.25 | 6.4x |

The difference was largest for omissions: clinician notes carried 7.3x the Critical+High omission density of AI notes under the calibrated reviewer (0.683 versus 0.094 per note) and were 5.2x as likely to carry at least one (40.8% versus 7.8%). This pattern was directionally consistent across error categories (Figure 3; Supplementary Figures S6 and S8): AI Critical+High errors were predominantly unsupported content — fabricated patient details, medications and examination findings — whereas clinician Critical+High errors were predominantly omissions: absent medication information, safety-netting and patient counselling. The omission difference persisted after adjusting for note length and was reproduced independently in the human-adjudication arm, which uses no automated reviewer.

**Figure 3.**
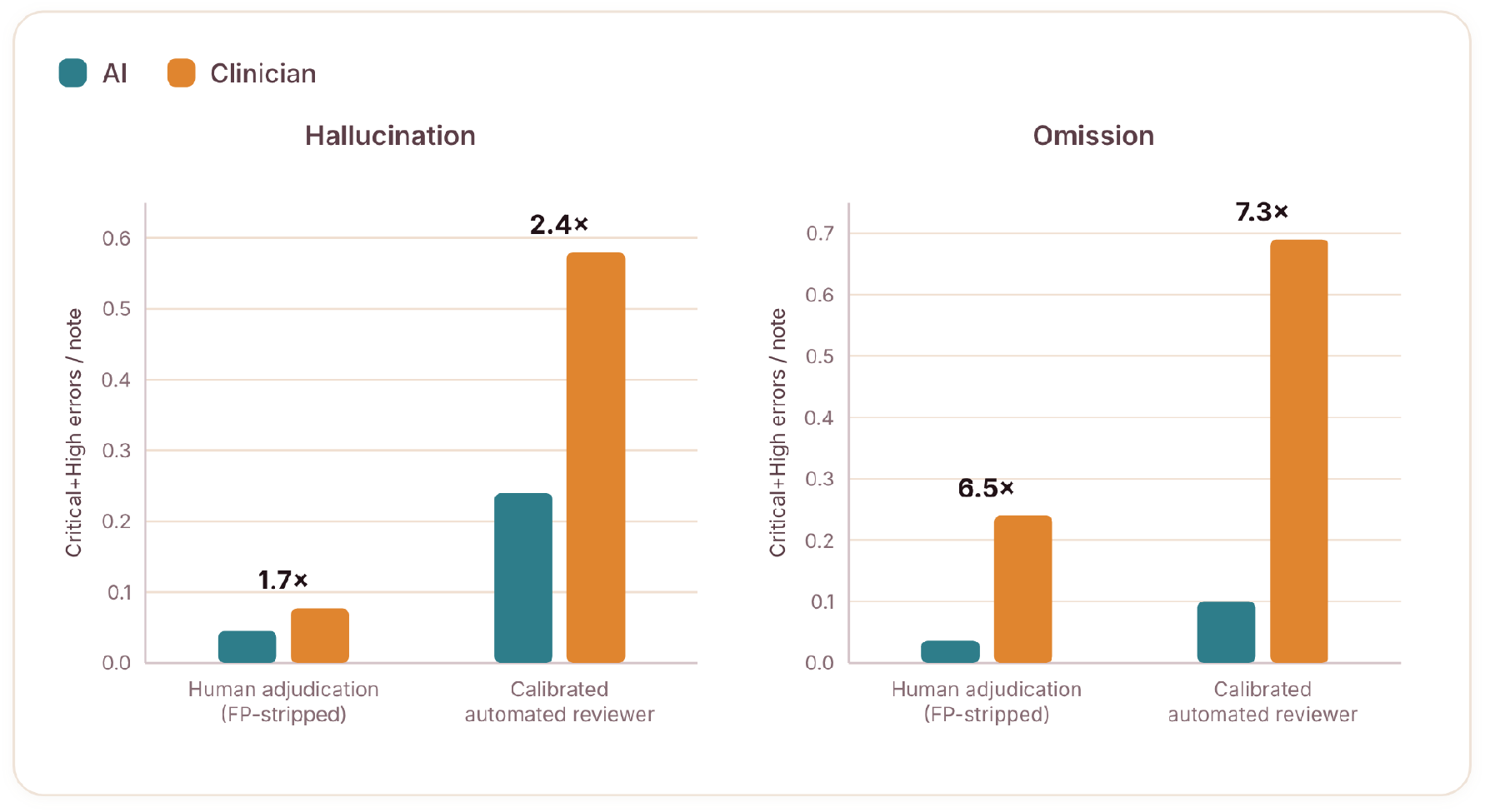
Critical+High errors per note, by error type and detection arm. Annotations give the clinician:AI ratio. The direction is the same under both detection arms and the difference is larger for omissions; the calibrated reviewer (right pair in each panel) identifies several-fold more errors than human adjudication (left pair) in both arms. Source: authors’ analysis of the 385-consultation paired dataset.

Benchmarked against the errors the automated reviewer confirmed, unaided clinician review identified only 11.5% of confirmed commission errors and 12.0% of confirmed omissions overall, and fewer in AI notes than in clinician notes (5.2% versus 14.0% for omissions; Supplementary Figure S9), consistent with an automation-bias mechanism in which fluent, well-structured prose makes residual errors harder for a human reader to notice.^35,36^ Because severity grading underpins the safety outcome, we examined inter-rater reliability of the three-model panel: pairwise tier judgements were within one adjacent tier in 89.4% of comparisons, and agreement on the Critical+High cutpoint was Gwet’s AC1^37^ 0.57. The unweighted four-tier Fleiss kappa^38^ was low (0.085), reflecting the kappa paradox under a heavily imbalanced marginal^39^ rather than an absence of agreement. Severity grading was performed blind to note authorship and identically across arms, so any residual grading noise is non-differential and biases the clinician-versus-AI contrast toward the null, meaning the observed safety advantage is conservative rather than inflated by grader unreliability.

Using a distribution-based MCID of 3.49 points, 54.3% of AI notes exceeded their paired clinician note by at least that margin. The Supplementary Appendix reports the component analyses: joint error profiles (Figure S6), restriction to the Critical tier (Figure S7), the Critical+High omission taxonomy with worked examples (Table S7), and the directional-bias checks confirming the safety result is not an artefact of sponsor-conducted grading (Table S3).

## Discussion

In this paired simulation across five languages and health systems, AI-generated notes scored higher on the PDQI-9, were less dispersed, and carried fewer Critical+High errors than clinician-written notes from the same consultations, consistently across sites, specialties, and noise conditions, and under both error-detection arms. Two features of the result matter more than the difference in means. First, dispersion: the arms differed nearly five-fold in the frequency of low-scoring notes (5.7% versus 27.8%). If low-quality documentation carries disproportionate downstream risk, this lower-tail difference may matter more to a health system than the shift in central tendency. Second, the largest difference we observed was in omissions rather than fabrications. A framing in which AI documentation risk is equated with fabrication risk, while clinician documentation is treated as the safe baseline, is not supported by these data.

The magnitude of the measured safety difference depends on the detector. The two arms differed several-fold in absolute error rates while agreeing on direction, echoing evidence that human record review detects only a minority of adverse events identifiable by systematic methods.^10^ This has a methodological implication for the field: studies relying on human adjudication alone will report low error rates for both AI and clinician notes, and may lack the sensitivity to detect a difference at all.^9,12^ We therefore report both regimes and bound the absolute rate rather than claiming a point estimate. Where the externally validated screen behaved asymmetrically, it did so against the direction we report: of the candidate flags the screen removed, 42% were confirmed by the external panel as genuine non-errors on clinician notes versus 56% on AI notes (Supplementary Table S3), meaning genuine clinician-note errors were discarded more often than genuine AI-note errors and the reported gap is, on this axis, conservative.

This study has several limitations. Blinding was to authorship labels, not authorship cues; AI notes were structurally distinctive, so functional unblinding cannot be excluded, though the transcript-grounded safety outcome does not depend on evaluator blinding. Documentation occurred in a simulated setting without the medicolegal accountability of real practice, and consultations were actor-performed from scripts rather than live encounters, yielding the known ground truth that error adjudication requires but not a full reproduction of live clinical practice. We assessed the AI’s unedited first draft; how much of this error burden survives clinician review before filing remains open. PDQI-9 inter-rater reliability was low in this cohort (Supplementary Table S4), which constrains absolute score interpretation more than the paired within-consultation contrast. The PDQI-9 was professionally translated for the four non-English sites, but formal language-specific psychometric validation is not available; the consistent direction of the paired effect across all five sites supports robustness of the comparative finding but does not substitute for such validation. The measurement instruments are proprietary, limiting independent re-implementation, although their behaviour was externally audited and the largest effect is reproduced by the tool-independent human arm. Finally, the comparator was the junior-to-middle-grade documentation workforce by design; findings should not be extrapolated to consultant-authored documentation, which this study did not sample. Full detail, including detection-sensitivity and reliability analyses, is in the Supplementary Appendix.

## Conclusion

In this paired, multi-country simulation with a pre-registered analysis plan, AI-generated clinical notes scored higher on documentation quality, showed less variability, and carried fewer clinically significant errors than notes written on the same consultations by junior-to-middle-grade clinicians, consistently across five languages. Unaided clinician review detected a smaller fraction of confirmed errors in AI-authored notes than in clinician-authored notes, which is relevant to any oversight model relying on unaided human review of generated documentation. Whether these findings extend to live clinical practice, consultant-authored documentation, and notes as filed after clinician editing remains to be established.

## Data Availability

Data supporting the findings of this study are reported in the article and accompanying Supplementary Materials. Additional study materials are provided where appropriate to support interpretation and reproducibility. The underlying evaluation datasets, source materials, and analysis or evaluation code may contain proprietary or commercially sensitive information and are not publicly available. Enquiries regarding access to additional materials may be directed to the corresponding author and will be considered subject to applicable confidentiality, intellectual-property, data-governance, and commercial requirements.

## Data and Code Availability

The de-identified session-level analysis dataset supporting the reported results (PDQI-9 total and item scores, error counts by class and severity tier, site, specialty, and acoustic condition) and the analysis code that reproduces every reported figure, table, and statistic from the frozen data snapshot are available on the Open Science Framework at https://osf.io/j46vz/overview (doi:10.17605/OSF.IO/J46VZ). The 299 Critical+High omission records underlying Supplementary Table S7, including assigned severity, are included in the same deposit.

Consultation scripts, full transcripts, and complete note texts are not publically released: the scripts are derived from real clinical encounters, and the note corpus constitutes an evaluation set for a commercial product. The error-identification judge, false-positive screen, and severity-grading harness are proprietary and are not released; their prompts and implementation are not published, and independent re-implementation of the detection pipeline is therefore not possible. Their behaviour was externally validated by a blinded panel of ten independent clinicians (Supplementary Table S3). Restricted materials may be made available to editors and peer reviewers under confidentiality agreement on request.

