## Supplementary Appendix for "Quality, consistency, and clinical safety of AI-generated versus clinician-written clinical notes: a multi-country paired simulation study"

This appendix contains the supplementary tables and figures referenced in the main text. Reference citations here are numbered independently of the main-text reference list and are listed in full at the end of this document.

### Table of Contents


| Item | Title |
|---|---|
| Table S1 | Cohort and participants by site |
| Table S2 | Departures from the registered analysis plan |
| Table S3 | Operating characteristics of the automated reviewer (external human validation) |
| Table S4 | Inter-rater reliability by site |
| Table S5 | Critical+High error density and burden by detection arm |
| Table S6 | Disposition of candidate flags by note authorship and error type |
| Table S7 | Critical+High omission taxonomy with worked examples |
| Table S8 | Allied Health arm, descriptive results |
| Table S9 | Bayesian model specification and convergence diagnostics |
| Figure S1 | Paired per-consultation outcomes on PDQI-9 |
| Figure S2 | Paired PDQI-9 distributions by site |
| Figure S3 | Per-writer mean PDQI-9 with within-writer standard deviation |
| Figure S4 | Item-level differences (AI minus clinician), PDQI-9 item means |
| Figure S5 | Random-effect standard deviations from the crossed model |
| Figure S6 | Joint error profile by note authorship |
| Figure S7 | Critical-tier burden and density |
| Figure S8 | Critical+High errors by category, by note authorship |
| Figure S9 | Error yield by detection arm, and human detection rate by note authorship |
| Figure S10 | AI quality is consistent across sites; clinician quality is not |
| File S1 | Registered statistical analysis plan (Open Science Framework) |

**Table S1. Cohort and participants by site.**

| Site | Paired consultations | Notewriters | Evaluators | Mean writer age (yr) | Sex recorded | Experience recorded |
|---|---|---|---|---|---|---|
| Cambridge, United Kingdom (English) | 80 | 8 | 20 | 28 | yes | yes |
| Barcelona, Spain (Spanish) | 79 | 8 | 10 | 29 | yes | yes |
| Milan, Italy (Italian) | 80 | 8 | 12 | 28 | yes | yes |
| Paris, France (French) | 80 | 8 | 14 | 29 | yes | yes |
| Cologne, Germany (German) | 66 | 8 | 11 | 30 | yes | yes |
| Pooled | 385 | 40 | 67 | 29 (24–34) | yes | 5/5 sites |

Note-writers were junior-to-middle-grade clinicians (Foundation Year to Specialty Registrar / international equivalents). Evaluators were blinded to note authorship (AI vs clinician). Eighty paired consultations were planned at each of the five sites (400 in total). The realised counts fall short of 80 at Barcelona (79) and Cologne (66) because of adjudicator non-completion: 15 sessions in total were left without evaluation data when adjudicators did not complete their assigned allocation within the field period. Non-completion was driven by adjudicator availability and scheduling and was unrelated to note content, note authorship or site performance, so the affected sessions are treated as missing completely at random and complete pairs only are analysed. No imputation was performed. Source: authors' analysis of the 385-consultation paired dataset.

**Table S2. Departures from the registered analysis plan.**

| Registered | As conducted | Effect on conclusions |
|---|---|---|
| Primary cross-site estimate to be the fixed effect of note_type from the mixed model; pooled unadjusted comparison "descriptive only" | Pooled paired Wilcoxon (+5.08) presented as the primary result, mixed model (−5.16) as corroboration | None; the two agree closely, but the reporting hierarchy is inverted relative to the plan |
| Rank-biserial correlation as the primary effect size, matched to the Wilcoxon test; Cohen's d secondary | Cohen's dz reported throughout; rank-biserial not reported | Direction and statistical significance unaffected: for a paired Wilcoxon test, Cohen's dz and the rank-biserial correlation order the arms identically. The substitution changes the scale on which the effect is expressed, not the inference. The registered primary effect size is not reported here. |
| Mixed model fitted by REML with ML for likelihood-ratio comparison; marginal and conditional $R^2$ reported | Bayesian crossed-random-effects model (bambi/PyMC); $R^2$ not reported | None expected; change of estimation framework, $R^2$ absent |
| ICC two-way mixed, absolute agreement | One-way random ICC(1), raters differing across notes by design | ICC(1) is the appropriate estimator given the realised rater assignment |
| Severity graded by two pre-specified LLM raters, mean of the two, with manual clinical adjudication of discordant cases | We used a three-model panel with majority-vote consensus. The manual clinical adjudication of discordant gradings was not carried out. | Material departure from a pre-specified procedure. The manual clinical adjudication of discordant severity gradings was not performed, so severity grading is wholly automated. The reliability of the three-model panel is reported in the main text (adjacent-tier agreement 89.4%; Gwet's AC1 0.57 on the Critical+High cutpoint), and grading was blind to authorship and identical across arms, so residual grading noise is non-differential and biases the |

| | | |
|---|---|---|
| | | AI-versus-clinician contrast toward the null. |
| Risk-matrix plot as the primary representation of risk | We report Critical+High density and burden as the primary safety summaries in place of the registered risk-matrix plot. | Substantive re-specification of the primary safety representation. Critical+High density and burden are reported in place of the registered risk-matrix plot; both are deterministic summaries of the same severity × likelihood scores, so no severity information is added or removed by the change of representation. |
| Qualitative review of the 10 highest-discrepancy sessions by two independent clinicians, blinded, with inter-reviewer agreement reported | We describe the three highest-discrepancy consultations narratively in the Discussion. The registered structured review by two independent blinded clinicians was not carried out. | Illustrative only; these consultations support no reported estimate. The registered structured review is not reported. |

Departures from the statistical analysis plan registered on the Open Science Framework (File S1) before pooling across sites. The registered plan is available at https://osf.io/j46vz (doi:10.17605/OSF.IO/J46VZ).

**Table S3. Operating characteristics of the automated reviewer against a blinded external clinician panel.**

Source: nested human-validation substudy.[1] Ten external clinicians (post-CCT GPs with emergency and surgical representation), 434 stratified pipeline flags, 565 adjudications, 131 items double-rated, English/Cambridge setting. Reviewers blinded to note authorship, identification source, pipeline verdict and severity tier.

**S3a. Agreement and concordance**

| Metric | Value | n | Interpretation |
|---|---|---|---|
| Inter-clinician agreement, genuine-error decision (raw) | 59% | 131 double-rated | No human consensus available as ground truth |
| Inter-clinician agreement, Gwet's AC1 | 0.24 | 131 | Fair |
| Judge–clinician agreement (raw) | 64% | 565 | Within the inter-clinician envelope |
| Pipeline concordance on clinician-consensus items | 70% | 77 | Consensus-genuine 68% (n=57); consensus-not-error 75% (n=20) |

**S3b. Non-differential behaviour across arms**

| Metric | AI notes | Clinician notes | Assessment |
|---|---|---|---|
| Human-confirmed precision, retained flags | 74% | 81% | Near-symmetric |
| Panel-vs-clinician signed severity gap (tiers) | +0.06 | −0.09 | Near-symmetric, non-directional |
| Removed flags confirmed as genuine non-errors | 56% | 42% | Asymmetric; screen over-removes genuine errors more often on clinician notes |

**S3c. Latent-class triangulation (Hui–Walter paradigm[2])**

Estimated within the flagged candidate pool only; not population error rates.

| Parameter | Posterior mean | 94% CI |
|---|---|---|
| Pipeline sensitivity | 0.83 | 0.68–0.98 |
| Pipeline specificity | 0.87 | 0.68–0.99 |
| Clinician sensitivity | 0.82 | 0.75–0.91 |

| | | |
|---|---|---|
| Clinician specificity | 0.57 | 0.45–0.75 |
| Genuine-error prevalence among flagged candidates | 0.68 | 0.48–0.83 |

**S3d. Severity-tier distribution, panel versus clinician**

Severity tiers follow the severity × likelihood risk matrix (range 1–25; Critical 16–25, High 10–15, Moderate 5–9, Low 1–4; higher = more severe), aligned with ISO 14971 medical-device risk management.[3]

| Tier band | LLM panel | Clinician |
|---|---|---|
| Critical/High | 28% | 24% |
| Moderate/Low | 72% | 76% |

n=309 genuine and graded. Gwet's AC1[4] is used as the prevalence-robust inter-rater agreement statistic in panel S3a. Source: nested human-validation substudy.[1]

**Table S4. Inter-rater reliability by site.**

| Site | Notes in analysis set | Notes double-rated | ICC (PDQI-9 total) | 95% CI | Mean evaluations per note | Notes with a single evaluation |
|---|---|---|---|---|---|---|
| Cambridge, United Kingdom (English) | 160 | 32 (20.0%) | 0.11 | −0.24 to 0.44 | 1.20 | 128 |
| Barcelona, Spain (Spanish) | 158 | 30 (19.0%) | 0.53 | 0.23 to 0.74 | 1.19 | 128 |
| Milan, Italy (Italian) | 160 | 32 (20.0%) | 0.45 | 0.12 to 0.68 | 1.20 | 128 |
| Paris, France (French) | 160 | 31 (19.4%) | 0.28 | −0.07 to 0.57 | 1.19 | 129 |
| Cologne, Germany (German) | 132 | 26 (19.7%) | −0.06 | −0.41 to 0.31 | 1.20 | 106 |
| Pooled | 770 | 151 (19.6%) | 0.24 | 0.09 to 0.39 | 1.20 | 619 |

PDQI-9 (Physician Documentation Quality Instrument): range 9–45, higher = better documentation quality; no validated minimal important difference has been published for this instrument.[5] ICC is the one-way random-effects ICC(1) on PDQI-9 total across the two independent ratings of each double-rated note (raters differ across notes by design).[6] No note received more than two independent evaluations. Source: authors' analysis of the 770-note paired dataset.

**Table S5. Critical+High error density and burden by detection arm.**

Calibrated automated-reviewer arm

| Error type | AI, per note | Clinician, per note | Clinician:AI ratio | Burden, AI | Burden, clinician |
|---|---|---|---|---|---|
| Commission — C+H density | 0.24 | 0.58 | 2.4x | 18.7% | 39.2% |
| Omission — C+H density | 0.094 | 0.683 | 7.3x | 7.8% | 40.8% |

Human-identification arm (FP-stripped)

| Error type | AI, per note | Clinician, per note | Clinician:AI ratio | Burden, AI | Burden, clinician |
|---|---|---|---|---|---|
| Commission — C+H density | 0.042 | 0.078 | 1.9x | 3.1% | 6.2% |
| Omission — C+H density | 0.039 | 0.25 | 6.4x | 3.1% | 17.1% |

Density = Critical+High errors per note, where Critical+High is the top two tiers of the severity × likelihood risk matrix (range 1–25, higher = more severe; Critical+High = 10–25), aligned with ISO 14971 medical-device risk management.[3] Burden = percentage of notes carrying at least one Critical+High error. 385 paired consultations (770 notes). The paired density and burden comparisons shown here are the pre-specified primary safety summaries; site-adjusted negative-binomial rate ratios with session pairing and note-writer clustering are not reported on this analysis set. Source: authors' analysis of the 385-consultation paired dataset.

**Table S6. Disposition of candidate flags by note authorship and error type.**

| Error type | Arm | Candidate flags | Retained as genuine | False positive | Reasonable inference | Uncertain |
|---|---|---|---|---|---|---|
| Commission | AI | 635 | 413 (65.0%) | 128 (20.2%) | 87 (13.7%) | 7 (1.1%) |
| Commission | Clinician | 971 | 801 (82.5%) | 51 (5.3%) | 105 (10.8%) | 14 (1.4%) |
| Omission | AI | 991 | 518 (52.3%) | 468 (47.2%) | — | 5 (0.5%) |
| Omission | Clinician | 3,019 | 1,956 (64.8%) | 1,060 (35.1%) | — | 3 (0.1%) |

Commission errors are statements in the note unsupported by, or contradicting, the consultation; omission errors are clinically relevant content present in the consultation but absent from the note.[7,8] For commission flags, false positive aggregates paraphrase supported, benign detail, and translation variant; for omission flags it aggregates appropriate concision, not clinically significant, and present but reworded. Flag dispositions are reported on the 400-consultation (800-note) pre-exclusion set rather than the 385-consultation analysis set, so the totals here exceed those implied by the main analysis (analysis-set flag totals: commission 616 AI and 936 clinician; omission 954 AI and 2,843 clinician). The disposition proportions, which are the quantity of interest in this table, are not materially affected, and all inferential safety results in the main text are computed on the 385-consultation analysis set. Source: authors' analysis of the calibrated automated-reviewer pipeline output.

**Table S7. Critical+High omission taxonomy with worked examples.**

Denominator: 299 omission flags graded Critical or High (severity × likelihood ≥10 on the 1–25 risk matrix[3]) that survived the false-positive screen, within the 385-consultation analysis set. Of the 299, 263 (88.0%) arose on clinician notes and 36 (12.0%) on AI notes; 27 were Critical and 272 High.

**S7a. Category distribution**

| Category | Definition | n | % of 299 | Critical | Clinician-note | AI-note |
|---|---|---|---|---|---|---|
| Medication information | Drug, dose, route, frequency or change discussed but absent or incompletely specified | 82 | 27.4% | 18 | 71 | 11 |
| Follow-up / disposition | Agreed review interval, referral, or conditional plan absent | 65 | 21.7% | 2 | 61 | 4 |
| Symptom / history documentation | Presenting symptom, past medical history, or substance/family/social history absent | 61 | 20.4% | 5 | 46 | 15 |
| Patient counselling | Material explanation, risk discussion or shared-decision content absent | 36 | 12.0% | 1 | 34 | 2 |
| Safety-netting | Explicit deterioration advice, red flags, or when to re-present absent | 30 | 10.0% | 0 | 29 | 1 |
| Objective findings | Vital signs, examination findings or results stated in the consultation absent | 24 | 8.0% | 1 | 21 | 3 |
| Allergy / adverse reaction | Stated allergy or prior adverse reaction absent | 1 | 0.3% | 0 | 1 | 0 |
| Total | | 299 | 100% | 27 | 263 | 36 |

**S7b. Worked examples**

Up to three per category (one for Allergy, the only such flag in the corpus), preferring Critical-tier flags and spanning the site-language cohorts and both arms. All transcript quotations were programmatically verified verbatim against the source consultation script. Non-English quotations are given in the original with an English gloss.

| Category | Site / arm | Transcript content (verbatim) | Absent from note | S×L | Tier |
|---|---|---|---|---|---|
| Medication information | Cologne, clinician | "…immer wenn ich das Mounjaro habe, setze ich es ganz niedrig, weil mein Zucker dann ständig abfällt.", with "ich hab's auf 120 gesetzt" | Note states no insulin for three months; current long-acting insulin dose (120) and the Mounjaro-linked adjustment pattern are absent | 20 | Critical |
| Medication information | Paris, clinician | "…laissez-moi vous donner quelque chose qui s'appelle le Meloxicam." | The prescribed drug is absent from the note entirely | 16 | Critical |
| Medication information | Milan, clinician | "…potassio 20 milliequivalenti una volta al giorno." | Absent from the home medication list | 16 | Critical |
| Medication information | Paris, AI | "Partons sur de l'amoxicilline, un comprimé trois fois par jour." | Note records corticosteroids and Mucinex but not the amoxicillin | 16 | Critical |
| Symptom / history documentation | Barcelona, clinician | "¿Pensamientos de que estaría mejor muerta… en las últimas dos semanas… ¿Entonces varios días? Sí." | The positive PHQ self-harm/suicidal-ideation item is absent | 20 | Critical |
| Symptom / history documentation | Barcelona, AI | "¿Pensamientos de que estaría mejor muerta… en las últimas dos semanas." | The positive PHQ-9 item 9 response is absent | 20 | Critical |
| Symptom / history documentation | Cambridge, clinician | "I've had two rounds of infusions, oral iron just doesn't really move my levels." | Note omits the failure of oral iron | 16 | Critical |
| Follow-up / disposition | Barcelona, clinician | "…tiene que traer el diazepam… lo vamos a desechar… y le recetaré los analgésicos." | Plan omits the surrender-before-prescribing condition | 16 | Critical |
| Follow-up / disposition | Cambridge, clinician | "…in about six months we'll bring you back in and look in your bladder again." | The planned repeat cystoscopy at six months is absent | 12 | High |
| Follow-up / disposition | Paris, clinician | "Pas de flexion, pas de torsion, pas de port de charge de plus d'un demi-kilo pendant les six premières semaines." | Note records only that activity advice was given | 12 | High |

| | | | | | |
|---|---|---|---|---|---|
| Patient counselling | Cambridge, clinician | “Those two medications together are dangerous, they both slow your central nervous system…” | Opioid-plus-benzo diazepine respiratory-depres sion counselling absent | 12 | High |
| Patient counselling | Milan, clinician | “…dovrà sospendere lo Zepbound un po’ prima dell’intervento… rischio di aspirare…” | Perioperative GLP-1 hold and aspiration-risk counselling absent | 16 | Critical |
| Patient counselling | Cambridge, AI | “With your lipoprotein A and the fact that you get migraines with aura…” | Oestrogen/contrac eption clotting-risk counselling absent | 12 | High |
| Safety-netting | Paris, clinician | “…vous devrez revenir si, une faiblesse d’un côté, un trouble de la parole… vous appelez le 15…” | No neurological red flags or emergency-call advice documented | 12 | High |
| Safety-netting | Cambridge, clinician | “…if you start having those thoughts… we will get off that medicine.” | Note omits stopping the medication as a safety-net action | 12 | High |
| Safety-netting | Cambridge, clinician | “You’d need to come back immediately if there was any more black stool, bright red blood, faintness, or very rapid pulse.” | Note omits bright red blood as a reason to return immediately | 12 | High |
| Objective findings | Milan, clinician | “C’è parecchio calcio nei vasi sanguigni intorno al cuore…” | No documentation of coronary calcium or calcium score | 16 | Critical |
| Objective findings | Cambridge, clinician | “Your oxygen level overnight has been holding at ninety-three, ninety-four percent on two litres of oxygen.” | Current saturation and oxygen requirement absent | 12 | High |
| Objective findings | Cologne, clinician | “Normales Kalzium liegt zwischen 8,5 und 10,8, Ihres ist 10,8; vorher 11,1…” | Hypercalcaemia values and trend absent | 12 | High |
| Allergy / adverse reaction | Cologne, clinician | “Und was passiert dann?”, “Ich kriege einen riesigen Ausschlag und schwelle an.” | Note lists the ibuprofen allergy but omits the nature of the reaction | 12 | High |

S×L: severity × likelihood score (range 1–25, higher = more severe); Tier: Critical (16–25) or High (10–15).[3] Source: authors' analysis of the 385-consultation paired dataset.

**Table S8. Allied Health arm, descriptive results.**

| Measure | AI | Clinician (allied-health professional) | Difference |
|---|---|---|---|
| n paired consultations | 15 | 15 | — |
| n notes | 15 | 15 | — |
| PDQI-9 mean (SD) | 37.3 (7.9) | 36.4 (7.2) | +0.87; Wilcoxon p=0.49; dz=0.15 |
| Errors per note (calibrated automated reviewer) | 3.93 | 9.33 | 2.4x |
| Critical+High commission density per note | 0.07 | 0.33 | 5.0x |
| Critical+High omission density per note | 0.07 | 0.53 | 8.0x |
| Critical+High density per note, all error types | 0.13 | 0.87 | 6.5x |
| Notes containing a Critical+High error | 13.3% (2/15) | 26.7% (4/15) | 2.0x |

Excluded from the main analysis under a pre-specified criterion (absence of the inter-rater cross-over substudy; structurally different note type). Three disciplines (occupational therapy, physiotherapy, speech and language therapy), five paired consultations each, clean-audio condition only. PDQI-9: range 9–45, higher = better.[5] Critical+High: severity × likelihood ≥10 on a 1–25 scale, higher = more severe.[3] All inferential statistics here are exploratory given the small sample. Source: authors' analysis of the Allied Health arm dataset.

**Table S9. Bayesian model specification and convergence diagnostics.**

| Component | Prior | Justification |
|---|---|---|
| Intercept | Normal(μ = 38.09, σ = 24.69) | Centred on the observed response mean; weakly informative on the 9–45 PDQI-9 scale |
| note_type coefficient (HU vs AI; HU = human, i.e. clinician-written) | Normal(μ = 0, σ = 34.92) | Mean-zero; σ far exceeds any plausible effect, so the posterior is data-dominated |
| SD, session random effect | HalfNormal(σ = 24.69) | Weakly-informative positive-support prior, scaled to the response |
| SD, evaluator random effect | HalfNormal(σ = 24.69) | As above |
| SD, note-writer random effect | HalfNormal(σ = 24.69) | As above |
| Residual SD | HalfStudentT(ν = 4, σ = 6.99) | Heavy-tailed, scaled to the residual SD of the response |

Model: PDQI-9 total[5] ~ note_type + (1|session_id) + (1|evaluator) + (1|notewriter), Gaussian family, identity link, fitted in bambi 0.18.0 on PyMC, where note_type contrasts clinician-written (HU) against AI notes. 770 observations; 385 sessions, 67 evaluator and 40 note-writer levels. Priors are bambi's default weakly-informative, data-scaled specification (response mean 38.09, SD 6.99).

| Parameter | Posterior mean | 94% HDI | R-hat | Bulk ESS | Tail ESS |
|---|---|---|---|---|---|
| Intercept (AI notes) | 40.61 | 40.0 to 42.0 | 1.00 | 2828 | 3853 |
| note_type[HU] (clinician-written) | −5.16 | −5.90 to −4.42 | 1.00 | 15255 | 5520 |
| SD, session | 0.50 | 0.03 to 1.3 | 1.00 | 2107 | 3221 |
| SD, evaluator | 3.43 | 2.8 to 4.2 | 1.00 | 2975 | 4482 |
| SD, note-writer | 1.33 | 0.71 to 2.0 | 1.00 | 2122 | 3439 |
| Residual SD | 5.35 | 5.1 to 5.6 | 1.00 | 8979 | 6065 |

Across all 498 monitored parameters: maximum R-hat 1.00, minimum bulk ESS 2107, minimum tail ESS 3221, 0 divergent transitions. Sampler: 4 chains, 2000 draws each after 2000 tuning steps, target_accept = 0.95, NUTS, random seed 20260518. Source: authors' analysis of the 770-note paired dataset.

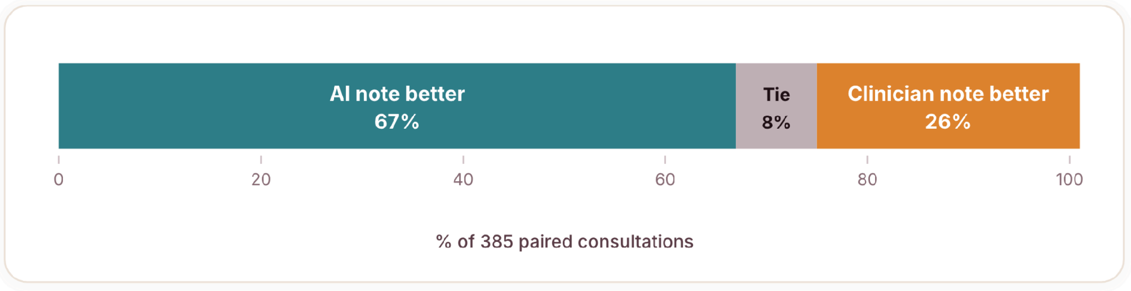


**Figure S1. Paired per-consultation outcomes on PDQI-9.** Percentage of 385 paired consultations in which the AI note scored higher (66.8%), equal (7.5%) or lower (25.7%) than its paired clinician note. PDQI-9: range 9–45, higher = better documentation quality; no validated minimal important difference has been published for this instrument.[5] Source: authors' analysis of the 385-consultation paired dataset.

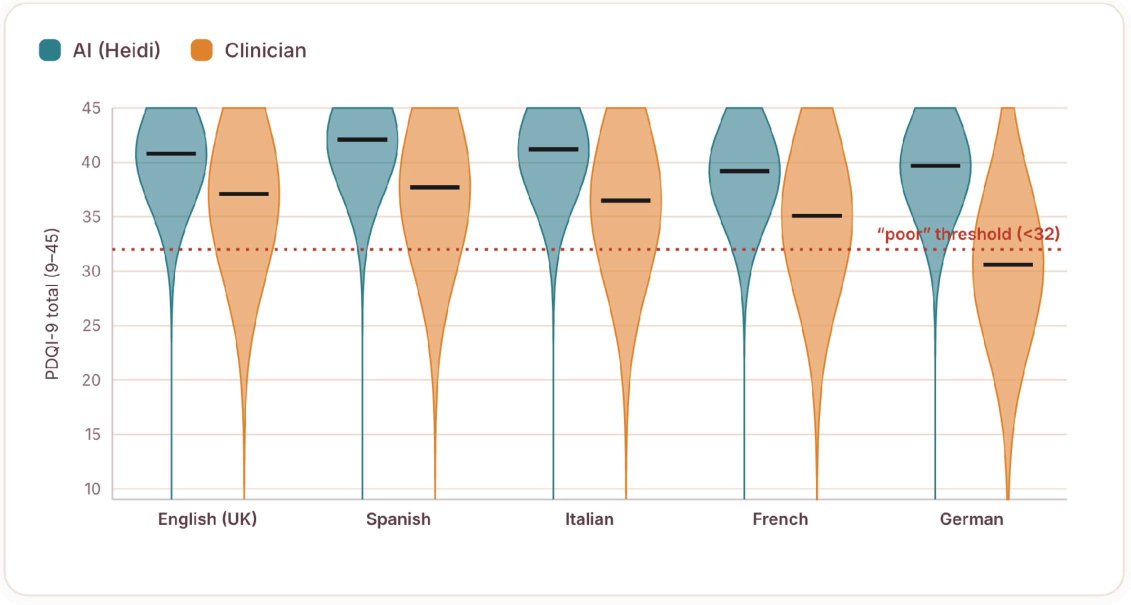


**Figure S2. Paired PDQI-9 distributions by site.** AI distributions are located higher on the scale and are narrower; clinician distributions are lower with a longer left tail extending below the pre-specified "poor" threshold (<32, dotted). The threshold was defined in the registered analysis plan as the 25th percentile of the pooled PDQI-9 distribution across both note types, and realised at 32 points on this dataset. It is therefore a distributional cut-point fixed before the arm-level results were inspected, adopted because no validated minimal important difference has been published for this instrument.[5] PDQI-9: range 9–45, higher = better documentation quality.[5] Source: authors' analysis of the 385-consultation paired dataset.

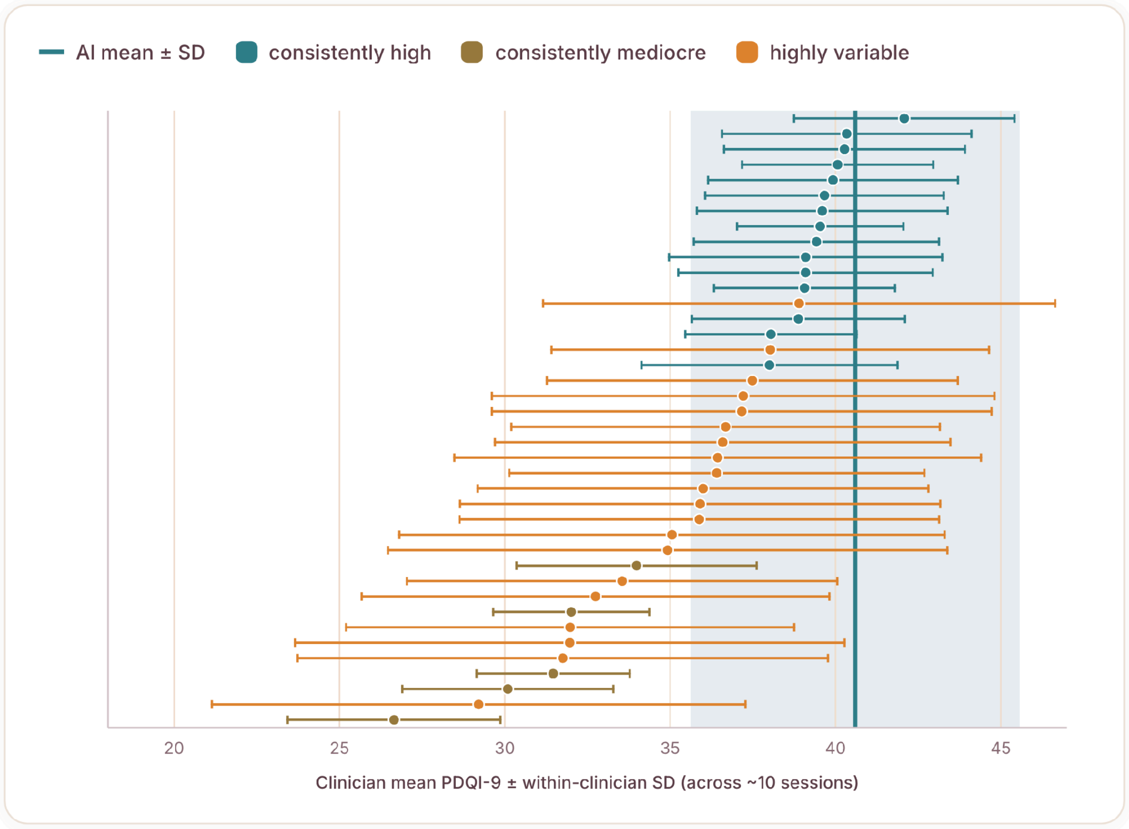


**Figure S3. Per-writer mean PDQI-9 with within-writer standard deviation.** Each row is one note-writer, sorted by mean, across their sessions; the shaded band and vertical line show the AI mean ± SD. PDQI-9: range 9–45, higher = better documentation quality.[5] Source: authors' analysis of the 385-consultation paired dataset.

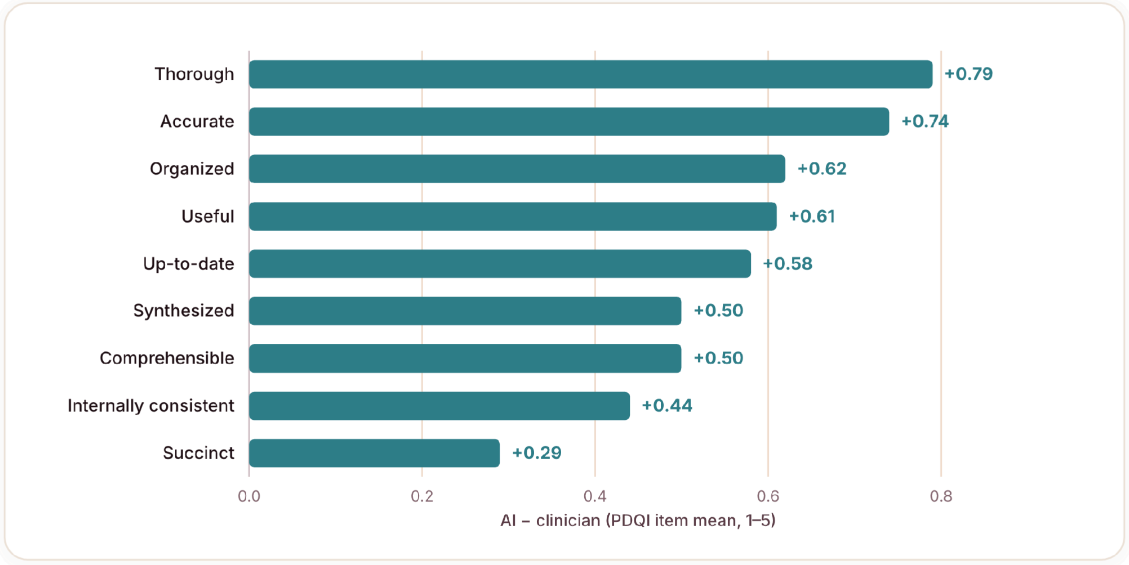

**Figure S4. Item-level differences (AI minus clinician), PDQI-9 item means.** All nine items favour the AI note; the largest differences are on Thorough, Accurate and Organized, and the smallest on Succinct. PDQI-9 item scale: range 1–5 per item, higher = better.[5] Source: authors' analysis of the 385-consultation paired dataset.

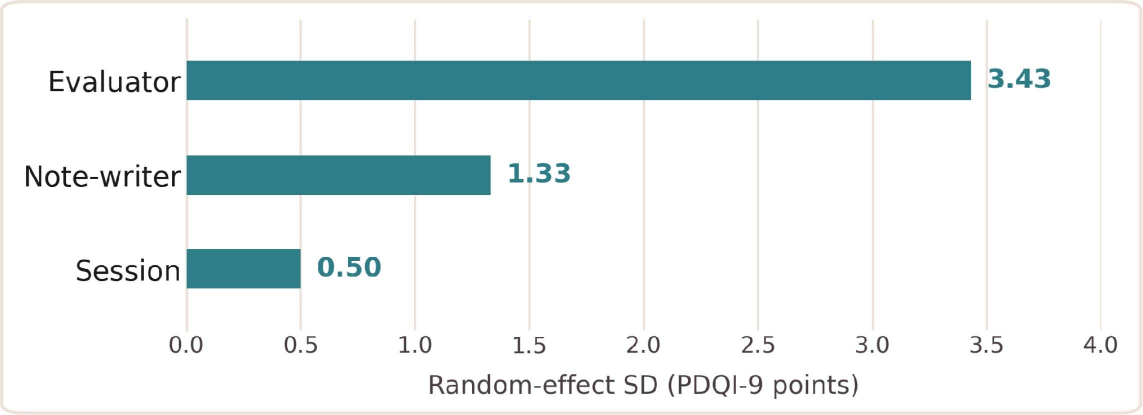


**Figure S5. Random-effect standard deviations from the crossed model.** Evaluator scoring tendency (3.43) exceeds both note-writer (1.33) and session (0.50) components. Units are PDQI-9 points (scale range 9–45, higher = better).[5] Source: authors' analysis of the 385-consultation paired dataset.

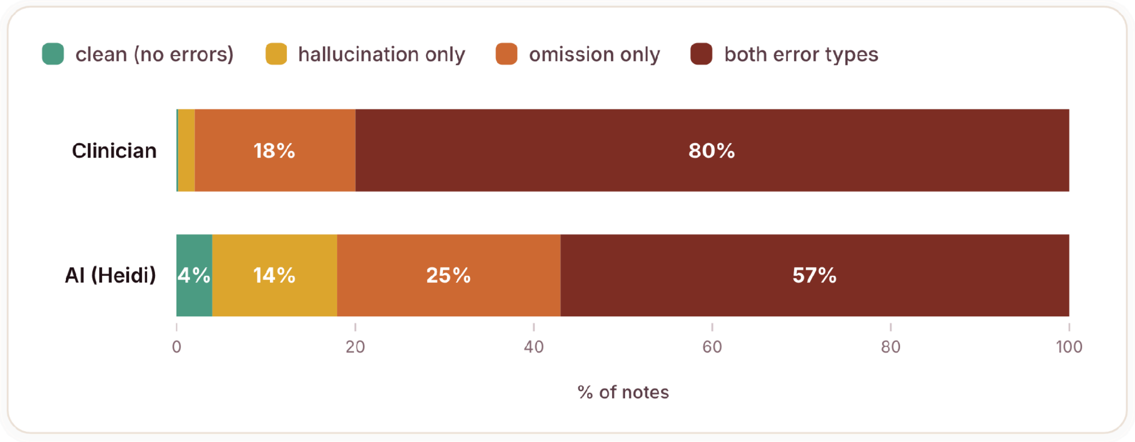


**Figure S6. Joint error profile by note authorship.** Percentage of notes classified as clean, commission-only, omission-only, or carrying both error types (all severities). Commission and omission are defined per the standard taxonomy for generated clinical text.[7,8] Source: authors' analysis of the 385-consultation paired dataset.

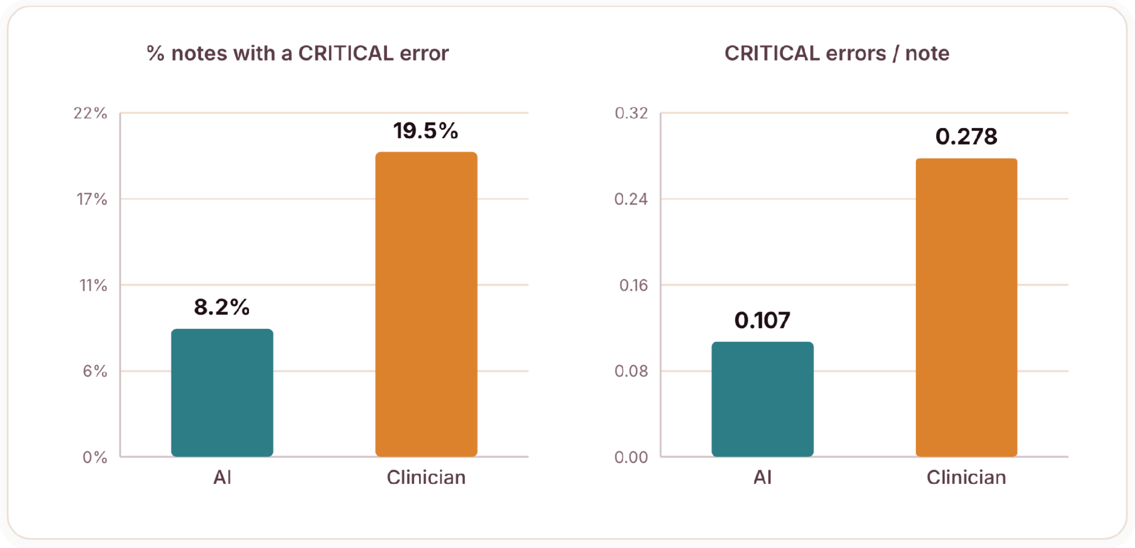


**Figure S7. Critical-tier burden and density.** The proportion of notes carrying any Critical error (left) and Critical errors per note (right). Critical: severity × likelihood ≥16 on a 1–25 risk matrix, higher = more severe.[3] Source: authors' analysis of the 385-consultation paired dataset.

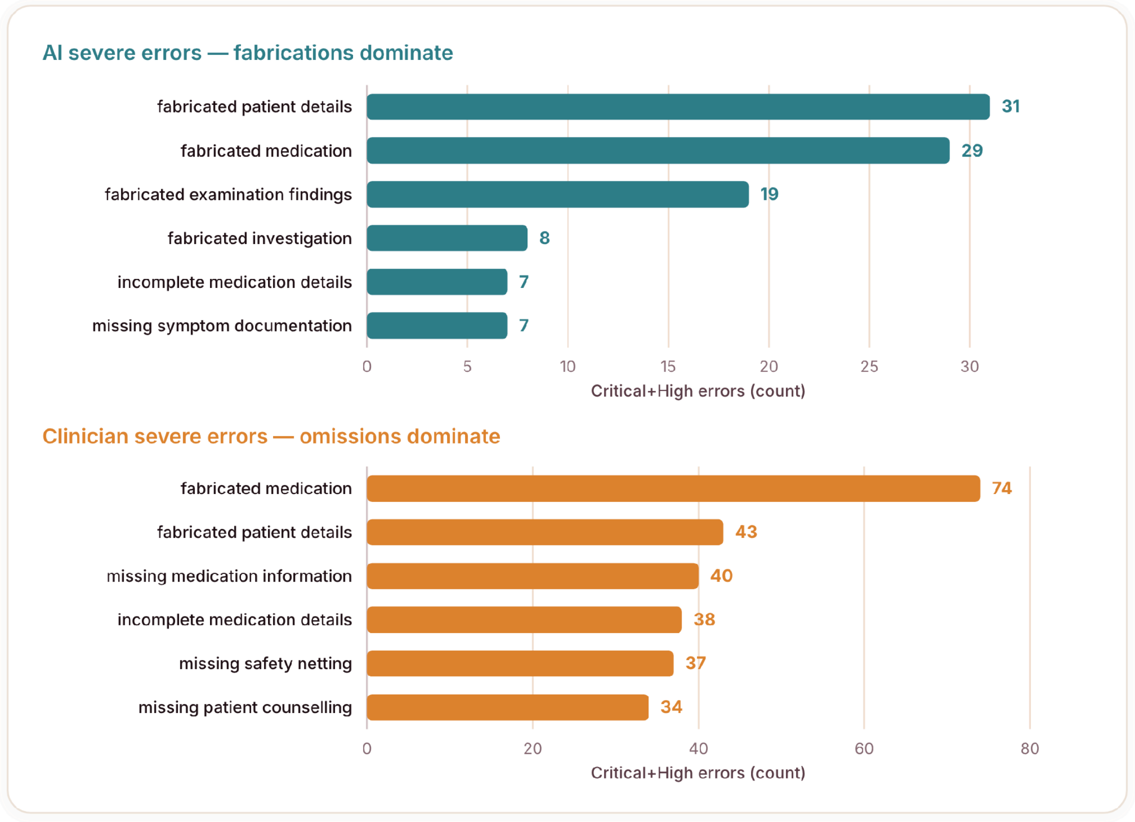


**Figure S8. Critical+High errors by category, by note authorship.** AI severe errors are concentrated in unsupported-content categories; clinician severe errors are concentrated in omission categories. Critical+High: severity × likelihood ≥10 on a 1–25 risk matrix, higher = more severe.[3] Source: authors' analysis of the 385-consultation paired dataset.

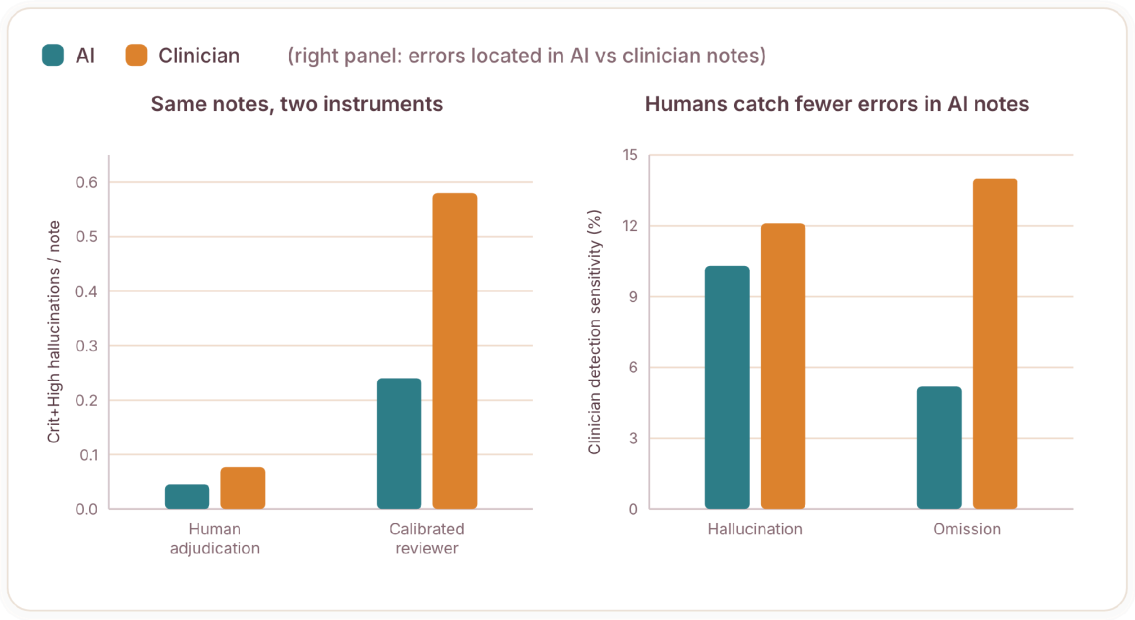


**Figure S9. Error yield by detection arm, and human detection rate by note authorship.** Left: Critical+High commission density in the same notes under the calibrated reviewer versus under human adjudication. Right: clinician detection rates are lower for errors in AI notes than in clinician notes. Critical+High: severity × likelihood ≥10 on a 1–25 risk matrix, higher = more severe.[3] Source: authors' analysis of the 385-consultation paired dataset.

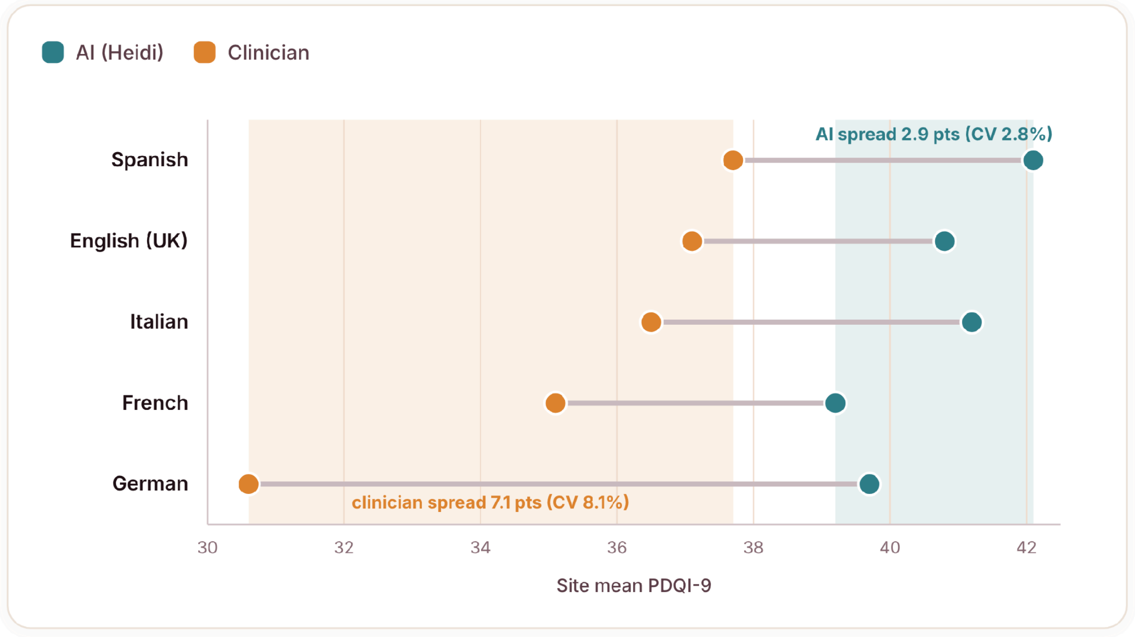


**Figure S10. AI quality is consistent across sites; clinician quality is not.** Site mean PDQI-9 by arm. Each dumbbell links a site's clinician mean to its AI mean. PDQI-9: range 9–45, higher = better documentation quality.[5] Source: authors' analysis of the 385-consultation paired dataset.
